# Unravelling Robust Brain-Behavior Links of Depressive Symptoms Through Granular Network Models: Understanding Heterogeneity and Clinical Implications

**DOI:** 10.1101/2023.09.13.23295278

**Authors:** René Freichel, Agatha Lenartowicz, Linda Douw, Johann D. Kruschwitz, Tobias Banaschewski, Gareth J. Barker, Arun L.W. Bokde, Sylvane Desrivières, Herta Flor, Antoine Grigis, Hugh Garavan, Andreas Heinz, Rüdiger Brühl, Jean-Luc Martinot, Marie-Laure Paillère Martinot, Eric Artiges, Frauke Nees, Dimitri Papadopoulos Orfanos, Tomáš Paus, Luise Poustka, Nathalie Holz, Christian Baeuchl, Michael N. Smolka, Nilakshi Vaidya, Robert Whelan, Vincent Frouin, Gunter Schumann, IMAGEN Consortium, Henrik Walter, Tessa F. Blanken

## Abstract

**Background:** Depressive symptoms are highly prevalent, present in heterogeneous symptom patterns, and share diverse neurobiological underpinnings. Understanding the links between psychopathological symptoms and biological factors is critical in elucidating its etiology and persistence. We aimed to evaluate the utility of using symptom-brain networks to parse the heterogeneity of depressive symptomatology in a large adolescent sample.

**Methods:** We used data from the third wave of the IMAGEN study, a multi-center panel cohort study involving 1,317 adolescents (52.49% female, mean±SD age=18.5±0.72). Two network models were estimated: one including an overall depressive symptom severity sum score based on the Adolescent Depression Rating Scale (ADRS), and one incorporating individual ADRS symptom/item scores. Both networks included measures of cortical thickness in several regions (insula, cingulate, mOFC, fusiform gyrus) and hippocampal volume derived from neuroimaging.

**Results:** The network based on individual symptom scores revealed associations between cortical thickness measures and specific symptoms, obscured when using an aggregate depression severity score. Notably, the insula’s cortical thickness showed negative associations with cognitive dysfunction (partial cor.=-0.15); the cingulate’s cortical thickness showed negative associations with feelings of worthlessness (partial cor. = -0.10), and mOFC was negatively associated with anhedonia (partial cor. = -0.05).

**Limitations:** This cross-sectional study included participants who were relatively healthy and relied on the self-reported assessment of depression symptoms.

**Conclusions:** This study showcases the utility of network models in parsing heterogeneity in depressive symptoms, linking individual symptoms to specific neural substrates. We outline the next steps to integrate neurobiological and cognitive markers to unravel MDD’s phenotypic heterogeneity.

## Introduction

Depressive symptoms continue to be highly prevalent across the globe, with increasing rates among adolescents and young people (Goodwin et al., 2022). Depression is a highly heterogeneous disorder (Goldberg, 2011) diagnosed based on the presence of five out of nine DSM-5 symptoms. These symptoms are however diverse, ranging from weight loss or gain to depressed mood, and contribute to disorder heterogeneity that poses challenges for treatment. Symptom network models have been used to capture this heterogeneous symptom expression as they conceptualize mental disorders as systems of interacting symptoms. The heterogeneity observed at the depression symptom level is intriguingly matched by the multifaceted neurobiological underpinnings of depression (Buch & Liston, 2021). Meta-analytical evidence points to neuroanatomical alterations in depression in adult samples, namely lower hippocampal volume (Schmaal et al., 2016) and lower cortical thickness in several regions, including the insula, cingulate, orbitofrontal cortex, and the fusiform gyrus (Schmaal et al., 2017). Modeling this interplay between symptom expression and biology is crucial for understanding depression’s etiology, and ultimately treatment (Remes et al., 2021).

However, when both domains (i.e., psychological/biological) are combined, then typically at least one domain is simplified in the process (Blanken et al., 2021), often to a single aggregate dimension. Most studies examining associations between structural and functional neural alterations and depressive symptoms, either use depression sum scores or subscales (aggregating the psychological level) or they use aggregate measures derived from neuroimaging, such as overall cortical thickness, or structural or functional connectivity, (aggregating the biological level). This abstraction potentially obscures more fine-grained associations, that could potentially account for the symptom heterogeneity.

One recent pilot study that included both brain and individual symptom measures into one network model did reveal cross-construct (i.e., brain-symptom) relations even in a small sample of depressed and never-depressed adults (Hilland et al., 2020). This finding suggests that fine-grained associations could indeed be obscured when using aggregate measures, but this was not evaluated directly. Network analysis (Borsboom et al., 2021) provides a suitable methodological approach for studying granular cross-construct associations between individual symptoms and brain markers considering 1) its focus on identifying partial associations (i.e., controlling for all other nodes) and 2) the use of regularization (i.e., shrinking weak estimates to zero) to lower the likelihood of false positive connections when estimating many parameters.

In the present study, we replicate the approach by Hilland et al. (2020) in a substantially larger sample to identify relations between depression symptoms and five a-priori selected (based on Hilland et al., 2020) brain markers (cortical thickness measures for insula, cingulate, mOFC, fusiform, and hippocampal volume). In addition, we will extend the previous study by directly evaluating whether parsing heterogeneity into individual symptom scores relative to an overall severity measure reveals cross-construct relations that otherwise would remain hidden.

## Methods

### Participants, procedure, and outcomes

We have used data from the third wave of IMAGEN study (Schumann et al., 2010), a multi-center panel cohort study of adolescents. Our final sample included 1,317 adolescents (52.49% female, M±SD = 18.5±0.7 years old) that completed the Adolescent Depression Rating Scale (ADRS) and 3D T1-weigthed gradient-echo Magnetic Resonance Imaging (MRI) scans. The ADRS is a validated 10-item self-report scale to assess the presence (present/ not present) of adolescent depression symptoms (Revah-Levy et al., 2007). The scale consists of 10 items that assess the presence of different depression symptoms on a binary scale (1 = *True/Present*, 0 = *False/Not present*). A total ADRS depression severity (sum) score above 6 is commonly used as a cut-off for a clinically relevant diagnosis of MDD as it ensures maximum sensitivity and specificity (Revah-Levy et al., 2007, 2011; Vulser et al., 2015).

The present sample showed substantial variability in the presence of all symptoms (see Table S1 in supplementary materials (SM) section 1), with 7% of individuals (*n=*89) meeting the criteria (score ≥ 6) for MDD. The magnetic resonance imaging (MRI) data was acquired using standard protocols to ensure homogeneity across scanners, including a 3D T1-weighted gradient echo volume (see SM1 and Schumann et al., 2010 for more details). Cortical thickness of insula, cingulate, mOFC, fusiform and hippocampal volume were estimated using the FreeSurfer software. We selected the same five brain regions (see Figure1) as Hilland et al. (2020) and followed their exact procedures: we averaged left/right hemispheres and used z-residuals for hippocampal volume (regressing out sex, intracranial volume). Age was not included as a covariate in the model considering that our sample was based on one assessment wave from a cohort study comprising adolescents of a comparable age group, with an average age of 18.5 years and a standard deviation of 0.7 years.

**Figure 1.**
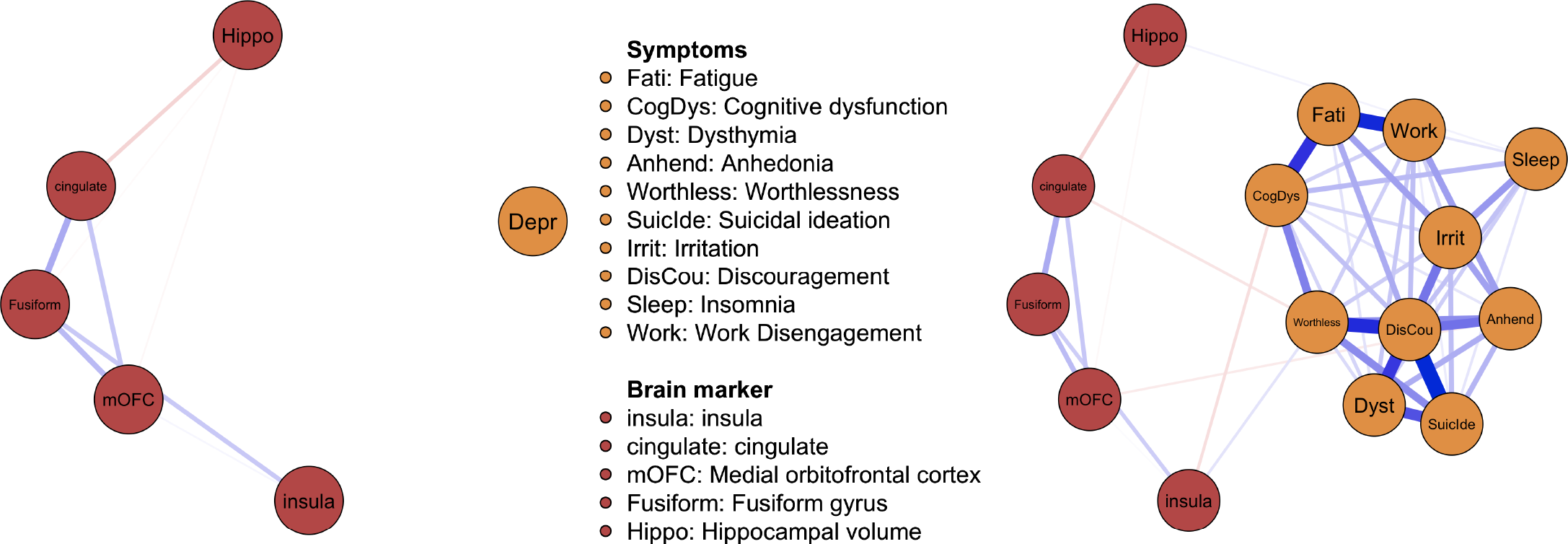
Depressive Symptoms – Brain Network Model. *Note*. The thickness of the lines indicates the strength of association. The connections (edges) in the network represent pairwise, partial associations between different symptoms and brain markers. Positive conditional associations are colored in blue, negative conditional associations are colored in red. Panel A includes the ADRS severity score (*Depr)*. Panel B includes all ADRS depression symptoms. The nodes for the brain regions (i.e., insula, cingulate, mOFC, Fusiform) refer to cortical thickness. *Hippo* refers to hippocampal volume. *mOFC* = medial orbitofrontal cortex. All edge weights can be found tabulated in supplementary Tables S2-S3.

### Statistical analysis

To investigate whether the abstraction of symptoms as sum scores obscures more fine-grained relations between brain regions and depression symptoms, we estimated two network models. Both networks contained the same brain measures (i.e., cortical thickness measures, hippocampal volume), however, one included the ADRDS sum score, indicating overall depression severity, and one included all individual ADRS items, representing different depression symptoms. We estimated both networks using LASSO regularization with cross-validation to minimize false positives (see SM2). We included the depression severity score as continuous and the single depression items as binary nodes (present/not present) included in the mixed graphical models (MGM, Haslbeck & Waldorp, 2020). The resulting connections in both networks (‘edges’) represent pairwise conditional associations (similar to partial correlations) that control for all other nodes in the network. While traditional statistical significance is not defined in these models, edges are included based on model fit. Included edges thus improve the fit of the model to the data. We assessed the edge weights’ accuracy using bootstrapping (*n*=1,000, see SM2).

## Results

The network including depression severity is shown in Figure 1A. We found no cross-construct associations between any of the neural markers and overall depression severity. In contrast, we found many positive associations within the respective domains (i.e., among depressive symptoms and cortical thickness measures) in the network estimated on the separate depression symptoms (Fig. 1B). The networks were sufficiently stable and all cross-modal links were retrieved in at least half of the bootstrapped samples (range 53-85%). Interestingly, we found cross-construct associations between cortical thickness measures and specific symptoms: cingulate was negatively associated with *worthlessness* (retrieved in 59%), insula was negatively associated with *cognitive dysfunction* (85% retrieved), and mOFC was negatively associated with *anhedonia* (53% retrieved). We found positive associations between insula and *worthlessness* (61% retrieved) and between hippocampal volume and *sleep* problems (60% retrieved).

## Discussion

The present study is one of the first to pinpoint granular associations between neural substrates of overall depressive symptomatology and specific depression symptoms using an integrated network approach. Crucially, we showed that these robust associations remain hidden when only including overall depression severity, concealing the heterogeneous symptoms. The negative associations shown (between regional cortical thickness and symptoms) align with prior work suggesting cortical thinning as a depression biomarker (Suh et al., 2019), in line with vulnerability theories of depression. Interestingly, our results also uncovered novel links, such as positive associations between *insula* and *worthlessness*.

We believe that our findings have dual implications; with respect to guiding future brain-behaviour research and bear relevance for clinical practice. First, our comparative analysis of networks estimated on an aggregate measure of depression severity (Fig. 1A) and specific depression symptoms (Fig. 1B) showed stark differences. The heterogeneity underlying the association between neural substrates and depressive symptoms was obscured when using an aggregate score. This suggests that networks estimated at the level of individual symptoms and neural makers have the potential to dissect these hidden associations and may allow us to better grasp the heterogeneity of depression.

Second, in addition to a better understanding of heterogeneity in psychopathology, symptom-brain networks, as showcased in this report, provide a better basis for discerning the clinical implications of these connections. For instance, in line with recent efforts in the field of precision psychiatry, brain-symptom networks may form the basis for insights relevant for the targeting of individual symptom – neural substrate connections (e.g., thinning of insula – cognitive dysfunction).

### Limitations

A limitation of our study is the self-reported assessment of depression symptoms that may naturally be biased. In addition, our sample was relatively healthy, and thus, the cross-modal links should be understood as associations describing how variability in depression symptoms are linked to variability in the selected brain markers. Lastly, the cross-sectional nature of our study precludes any conclusions about the directionality and causal nature of the associations between neural markers and depression symptoms. The present study serves as a ‘proof-of-principle’ that may inspire future work to validate the mapping of symptoms and neural markers in clinical samples.

## Conclusions

Altogether, this brief report showcases the utility of brain-symptoms in the case of depression. Moving forward, future research should adopt such approaches and integrate neurobiological and cognitive markers to parse the phenotypic heterogeneity of depressive symptomatology both at a cross-sectional and developmental level.

## Data Sharing Statement

The data for this study are available from the IMAGEN study. Restrictions apply to the availability of these data, which were used under license for this study.

## Acknowledgements

This work received support from the following sources: the European Union-funded FP6 Integrated Project IMAGEN (Reinforcement-related behaviour in normal brain function and psychopathology) (LSHM-CT-2007-037286), the Horizon 2020 funded ERC Advanced Grant ‘STRATIFY’ (Brain network based stratification of reinforcement-related disorders) (695313), Human Brain Project (HBP SGA 2, 785907, and HBP SGA 3, 945539), the Medical Research Council Grant ‘c-VEDA’ (Consortium on Vulnerability to Externalizing Disorders and Addictions) (MR/N000390/1), the National Institute of Health (NIH) (R01DA049238, A decentralized macro and micro gene-by-environment interaction analysis of substance use behavior and its brain biomarkers), the National Institute for Health Research (NIHR) Biomedical Research Centre at South London and Maudsley NHS Foundation Trust and King’s College London, the Bundesministeriumfür Bildung und Forschung (BMBF grants 01GS08152; 01EV0711; Forschungsnetz AERIAL 01EE1406A, 01EE1406B; Forschungsnetz IMAC-Mind 01GL1745B), the Deutsche Forschungsgemeinschaft (DFG grants SM 80/7-2, SFB 940, TRR 265, NE 1383/14-1), the Medical Research Foundation and Medical Research Council (grants MR/R00465X/1 and MR/S020306/1), the National Institutes of Health (NIH) funded ENIGMA (grants 5U54EB020403-05 and 1R56AG058854-01), NSFC grant 82150710554 and European Union funded project ‘environMENTAL’, grant no: 101057429. Further support was provided by grants from: - the ANR (ANR-12-SAMA-0004, AAPG2019 - GeBra), the Eranet Neuron (AF12-NEUR0008-01 - WM2NA; and ANR-18-NEUR00002-01 - ADORe), the Fondation de France (00081242), the Fondation pour la Recherche Médicale (DPA20140629802), the Mission Interministérielle de Lutte-contre-les-Drogues-et-les-Conduites-Addictives (MILDECA), the Assistance-Publique-Hôpitaux-de-Paris and INSERM (interface grant), Paris Sud University IDEX 2012, the Fondation de l’Avenir (grant AP-RM-17-013), the Fédération pour la Recherche sur le Cerveau; the National Institutes of Health, Science Foundation Ireland (16/ERCD/3797), U.S.A. (Axon, Testosterone and Mental Health during Adolescence; RO1 MH085772-01A1) and by NIH Consortium grant U54 EB020403, supported by a cross-NIH alliance that funds Big Data to Knowledge Centres of Excellence. This study is part of the project ‘New Science of Mental Disorders’ (www.nsmd.eu), supported by the Dutch Research Council and the Dutch Ministry of Education, Culture and Science (NWO gravitation grant number 024.004.016). T.F.B. was supported by an Amsterdam Brain and Cognition Project Grant.

